# DEFINE(prehospital): A protocol for a qualitative interview study to determine the pathway and response to the inclusion of frailty in pre-hospital trauma triage tools

**DOI:** 10.1101/2025.06.28.24307197

**Authors:** Philip Braude, Kim Kirby, Adam Bedson, Helen Nicholson, Jonathan Benger, Edd Carlton, Sarah Black, Joshua Chambers

**Affiliations:** Academic Geriatrician, North Bristol Trust and University of West of England; Senior Research Fellow Centre for Health and Clinical Research The University West of England Bristol; Research Paramedic, South Western Ambulance Service NHS Foundation Trust; Senior Research Fellow, Centre for Health and Clinical Research, The University West of England Bristol; Professor of Emergency Care, University of the West of England Bristol; Chief Medical Officer for NHS Digital; Consultant in Emergency Medicine, University Hospitals Bristol NHS Foundation Trust; NIHR Senior Investigator; Academic Emergency Medicine, University of Bristol, NIHR Advanced Fellow; Head of Research, Audit and Quality Improvement, South Western Ambulance Service NHS Foundation Trust; Clinical Fellow, North Bristol Trust

**Keywords:** Frailty, paramedics, trauma, triage Study Flow Chart

## Abstract

At the start of the COVID-19 pandemic South Western Ambulance Service Foundation Trust (SWASFT) changed its pre-hospital trauma triage tool to include an assessment of frailty, and if scored 5 or more on the Clinical Frailty Scale, the tool instructed the person to be conveyed to a trauma unit and not the major trauma centre. How the use of frailty has been operationalised and effected in daily clinical practice is unknown. There were concerns during the COVID-19 pandemic that care was being rationed by a frailty score alone, without expert oversight or appropriate training. Whether or not care rationing was appropriate at the height of the pandemic remains a philosophical question, however more research is needed into using the CFS for this type of decision making. This is relevant to using frailty scores in trauma triage tools outside of a pandemic situation, particularly when a score may prompt a change in clinical management.

This study aims to explore the pathways of care that have been associated with this incorporation of frailty. In addition, it will aim to explore healthcare professionals experience of frailty scoring for trauma patients pre-hospital and at the receiving hospitals.

## 1 Background

### Brief Study Description

Older people represent the largest group of patients in major trauma, with fall from standing being the most common mechanism of injury. Living with frailty is one factor that contributes to both presentations of older people with injuries and leads to worse outcomes compared to fitter patients. Using the Clinical Frailty Scale (CFS), Frailty is an independent predictor of 30-day mortality, inpatient delirium, and increased care level at discharge in older people admitted with trauma(1). However, it is not known how frailty has been used in the pre-hospital triage of older people, particularly during a d after the covid-19 pandemic.

Current geriatric-specific criteria used in pre-hospital trauma triage lacked accuracy, and patients had an increased chance of under-triage when they met the criteria (2). Under-triage, defined as transfer to lower-level trauma units or other acute care facilitates, as opposed to major trauma centers, of older adults with trauma has been demonstrated to be a problem. Trauma triage tools tend to be designed for the assessment of high impact trauma without any confounding by frailty or multimorbidity – potentially making them less accurate and relevant for older people living with frailty. Although there is currently no strong consensus surrounding the benefits of MTCs for patients with frailty, it is worth noting that most studies show at least a small benefit of MTC access for older adults(3)(4)(5), suggesting that diversion away due to age or frailty may not be supported by evidence.

However, there is limited evidence on the validity and feasibility of using frailty scores in trauma triage, in contrast to inpatient acute care where the CFS is now widely implemented. A qualitative interview study of paramedics exploring perceptions of frailty noted that there was variable knowledge and confidence in assessing for frailty. Although this paper was not trauma specific, themes arose about how frailty scores may influence ongoing care such as discharge planning, and access to multidisciplinary frailty-based services (6).

## 2 Rationale

At the start of the COVID-19 pandemic South Western Ambulance Service Foundation Trust (SWASF) changed its pre-hospital trauma triage tool to include an assessment of frailty, and if scored 5 or mo e on the clinical frailty scale, the tool instructed the person to be conveyed to a trauma unit and not the major trauma centre. How the use of frailty has been operationalised and effected in daily clinical practice is unknown. There were concerns during the COVID-19 pandemic that care was being rationed by a frailty score alone, without expert oversight or appropriate training(7). Whether or not care rationing was appropriate at the height of the pandemic remains a philosophical question, however more research is needed into using the CFS for this type of decision making. This is relevant to using frailty scores in trauma triage tools outside of a pandemic situation, particularly when a score may prompt a change in clinical management.

## 3 Theoretical Framework

This two-part qualitative study will aim to develop an understanding of how frailty has been incorporated within the pre-hospital environment for trauma:

1. Pathway mapping will allow a visualization of protocolized care, as well as mapping stakeholders’ experience and deviation from these pathways.
2. Reflexive thematic analysis of semi-structured interviews will be used to draw out the common themes from the interviews. This will allow this study to unpack the attitudes and experiences of healthcare professionals to act as a framework for future studies.

This preliminary work will inform three future studies:

1. Exploring patients’ and carers’ perception of the use of frailty in the pre-hospital trauma environment.
2. Exploration of other settings where frailty has been incorporated into trauma triage tools. Two other trauma triage tools are known of in other ambulance trusts, but both call for a discussion with a Senior Clinical Advisors if frailty is present, rather than a direct management change associated with a frailty score as in the SWASFT tool.
3. Consensus work around the use of frailty to ensure the provision of appropriate access to services and evidence based shared decision making.

## 4 Research Question

### 4.1 Objectives

1. To document pathways associated with the inclusion of a frailty assessment in pre-hospital trauma triage tools.
2. What have been responses of healthcare professional stakeholders to the inclusion of frailty assessment in pre-hospital trauma triage tools:
  a. To explore experiences of using frailty assessment in the pre-hospital setting
  b. To explore the experienced clinical response from hospitals to pre-hospital frailty assessment
  c. To compare the experienced clinical response in trauma units and the major trauma centre in relation to frailty assessment
  d. To explore views on the benefit and risks associated with frailty assessment in the prehospital setting
  e. To explore the facilitators and barriers to frailty assessment in the pre-hospital setting for trauma

### 4.2 Outcomes

1. Pathway map of the use of frailty surrounding the SWASFT trauma triage tool
2. Deviations from the pathway map
3. Common themes and examples from stakeholders in response to frailty scoring in pre-hospital trauma triage
4. Targets for improving the experience of staff using frailty scoring in trauma
5. Themes and questions to explore with patient and carers in future studies
6. Targets for future research around ensuring equitable patient access and transparency in the use of frailty

## 5Study Design and Methods of Data Collection

### 5.1 Study design

A qualitative study using semi-structured interviews and thematic analysis.

### 5.2 Data Collection

Sampling and recruitment are described in Section 7. Data will be collected using semi-structured interviews. Interviews will be performed by the Chief Investigator (PB) and recorded on Microsoft Teams. Interviews will be expected to take around 30-40 minutes, but have no maximum time limit. The topic guide will be developed by the Study Management Group during the study set-up phase with input from patient involvement representatives.

Anonymised transcripts of the interviews will be made by an independent transcriber.

### 5.3 Data Storage

Audio data will be recorded on Microsoft Teams using a secure North Bristol Trust server. Access to the data will be by email invitation. Data will be accessible only to the interview, interviewer, and University of West of England approved transcriber.

Transcriptions will be kept on North Bristol Trust secure servers and only accessible via password protected files. Data will be kept for 3 years after the study close in case of external requests to verify published material. Participants’ contact details will be stored separately from any notes, documents, transcripts and audio recordings.

### 5.4 Data Analysis

#### 5.4.1 Pathway Mapping

Stakeholders will be asked to describe the clinical pathway as well as give their experience of the pathway in operation. Microsoft Visio or PowerPoint will be used to develop a pathway map including deviations from the protocolized clinical pathway. Stakeholders will be asked to review the pathway map after each interview to ensure accuracy. This will occur up until closing of the study.

#### 5.4.2 Qualitative Interviews

Once transcribed, a well-established iterative process of data reduction, constant comparison, organisation and understanding through thematic analysis will be used:

- Familiarisation with the data: reading and re-reading transcripts
- Generating initial codes: noting codes of interesting and pertinent ideas
- Searching for themes: systematically organising these recurrent ideas with extracts of text
- Reviewing themes: checking themes are meaningful and relate to the text
- Defining and naming themes: summarising the narrative with clear definitions
- Producing the report: using extracts of data to exemplify the themes
- Each of these data sets will be analysed by the primary researcher with a sample checked by a second researcher for plausibility and validity. Consensus of themes will need to be reached (8).

## 6 Study Setting

The study will take place with stakeholders operating within the Severn Major Trauma Network. The area covers 1.2 million people in the South West of England. The major trauma centre is Southmead hospital, North Bristol Trust. Seven trauma units feed into the major trauma centre.

Participants will not be required to be in a particular location while the interview takes place as they will be conducted via video call.

## 7 Sample and Recruitment

### 7.1.1 Inclusion criteria

‐ Healthcare professionals:

○ employed by the NHS
○ operating within the Severn Major Trauma Network
○ expected to include, but not limited to:

▪ emergency department staff: doctors, nurses
▪ senior clinical advisors
▪ paramedics
▪ management
○ able to provide informed consent

### 7.1.2 Exclusion criteria

‐ Unable to provide informed consent
‐ Qualified at least 2 years (prior to the inclusion of frailty in the SWASFT tool)

## 7.2 Sampling

### 7.2.1 Size of sample

We will aim to recruit 8-12 participants. This is based on the breadth of stakeholders roles, rather than aiming to achieve saturation.

### 7.2.3 Sampling technique

We will use snowball sampling initially to identify potential participants. The study team and study participants will be encouraged to identify potential participants. This will allow a network of stakeholders to be identified who may have experience in the pre-hospital trauma pathway. It will allow identification of stakeholders that may be hidden to the study team.

Purposive sampling will be used to ensure diversity of the stakeholders to include a spectrum of job roles, gender balance, and years of seniority or years of experience in the field.

## 7.3 Recruitment

### 7.3.1 Sample identification

Initially key people known to the study team will be targeted for recruitment. The study team and study participants will be encouraged to identify potential participants. The study team will send details to the potential participant to see whether they would wish to be involved, and if so their details will be passed to the CI to send participant information sheet and consent form.

We will look to advertise the study in the SWASFT weekly bulletin and via social media (e.g. Twitter “Do you work in @SWASFT? We would like to interview you about how #frailty is used in the care of injured people. Please contact @DrPhilipBraude or email philip DOT braude AT nbt.nhs.uk”. This will aim to minimize bias to avoid a narrow range of views from participants that are known to each other. Interested potential participants will be asked to contact a central email address or Twitter handle direct message for more details. The CI will send participant information sheet and consent form Stakeholders will be invited to take part usually via Trust email systems. Eligibility criteria will be made clear in recruitment materials and confirmed by email before interviews are conducted.

### 7.3.2 Consent

The participant will be given 5 days to read the information before being re-contacted to answer any questions and ask if they are happy to participate. If content, the participant can return the consent form to the research team via secure email. Verbal consent will also be asked for before the start of each interview. The researcher will then contact the participant by email to explain the study procedures, answer any questions, and arrange a date and time for the first interview.

It will be made clear that healthcare professionals are under no obligation to take part. They will be free to withdraw at any time from the study up until data is anonymized

## 8 Ethical and Regulatory Considerations

### 8.1 Assessment and management of risk

It is not anticipated that the interviews will pose any significant risks to the participants. However, if they feel they are in any way unable or unwilling to continue with the interview and would like their data to be excluded from the study, they are able to stop the interview and withhold their data at any time without the need to offer an explanation. However, after the data has been anonymised it cannot be withdrawn.

It is possible that participating in interviews could lead to emotional distress. A risk assessment, detailing appropriate control measures, will be completed by the Study Management Group and logged by the Sponsor prior to recruitment and data collection taking place. If required we will offer participants a referral to their local health and wellbeing service.

### 8.2 Research Ethics Committee (REC) and other Regulatory review

HRA approval will be gained. A REC decision will not be sought in line with the HRA decision aid: “Under paragraph 2.3.14 of GAfREC, review by a REC within the UK Health Departments’ Research Ethics Service is not normally required for research involving healthcare or social care staff recruited as research participants by virtue of their professional role”. The study will also be reviewed by the South Western Ambulance Service NHS Foundation Trust Research group, and the University of the West of England ethics board. No research activities will begin until all research approvals are obtained.

#### Regulatory Review and Compliance

Before any site can enroll patients into the study, the Chief Investigator/Principal Investigator or designee will ensure that appropriate approvals from participating organisations are in place. Specific arrangements on how to gain approval from participating organisations are in place and comply with the relevant guidance.

For any amendment to the study, the Chief Investigator or designee, in agreement with the sponsor will submit information to the appropriate body for them to issue approval for the amendment. The Chief Investigator or designee will work with sites (R&D departments at NHS sites as well as the study delivery team) so they can put the necessary arrangements in place to implement the amendment to confirm their support for the study as amended.

#### Amendments

Any study amendments will be submitted for review by the study sponsor before submission to the HRA.

### 8.3 Peer review

The protocol has been reviewed by the Chief Investigator and study management group prior to being sent for assesssment by the nominated Clinical Trials Officer in the Research and Innovation Department at North Bristol Trust.

### 8.4 Patient Involvement

Three patients from the Department of Medicine for the Older Person’s patient feedback group were invited to contribute to work on frailty. Three members were interviewed focusing on studies and frailty.

Further patient involvement will take place around the interpretation and write-up of the study, as well as deciding on future steps.

### 8.5 Protocol Compliance

Accidental protocol deviations can happen at any time. They must be adequately documented on the relevant forms and reported to the Chief Investigator and Sponsor immediately.

Deviations from the protocol which are found to frequently recur are not acceptable, will require immediate action and could potentially be classified as a serious breach.

### 8.6 Data Protection and Patient Confidentiality

Individual participants are not being studied in this project. Once transcribed all participants data will be anonymized. However, there is potential for identification of participants given the key position held by them. This will be made clear in the consent from, although no names or titles will be used.

Interviews will be recorded on an encrypted device, anonymised and transcribed verbatim. The data will be anonymised using a PIN (Participant Identification Number) generated specifically for this study by the study coordinator.

It is not anticipated that the interviews will pose any significant risks to the participants. However, if they feel they are in any way unable or unwilling to continue with the interview and would like their data to be excluded from the study, they are able to stop the interview and withhold their data at any time without the need to offer an explanation. However, after the data has been anonymised it cannot be withdrawn. It is possible that participating in interviews could lead to emotional distress. A risk assessment, detailing appropriate control measures, will be completed by the Study Management Group and logged by the Sponsor prior to recruitment and data collection taking place.

Any notes, documents, audio-recordings and information about participants will be kept in the strictest confidence and managed in accordance with GDPR. Participants’ personal information will be stored separately from any notes, documents, transcripts and audio-recordings. Identifiable information will be securely erased on completion of the study.

### 8.7 Indemnity

This is an NHS-sponsored research study. For NHS sponsored research HSG(96)48 reference no.2 refers. If there is negligent harm during the clinical trial when the NHS body owes a duty of care to the person harmed, NHS indemnity covers NHS staff, medical academic staff with honorary contracts, and those conducting the trial. NHS indemnity does not offer no-fault compensation and is unable to agree in advance to pay compensation for non-negligent harm

### 8.8 Access to the Final Study Dataset

The anonymized data will only be available to the Chief Investigator (Philip Braude) and the study management group named in the protocol. Responses will be anonymised. Files will be password protected and stored on the hospital’s secure server, only accessible from a Trust computer with employee login. Any transfer of data required, e.g. from the approved transcription service, will occur only using secure servers and encrypted files.

In line with many peer reviewed journal’s policy for data sharing, data sharing may be offered to third parties only on request to the study CI with review by the study sponsor to ensure legitimate academic interest. Data will be shared with anonymous records if deemed appropriate, arranged via data sharing agreement, and transferred using secure systems. If requests originate from outside the EU this will be discussed with the study sponsor.

## 9 Dissemination Policy

### 9.1 Dissemination policy

The data will be owned by North Bristol Trust as the sponsor. On completion of the study the data will be analysed and a study report completed.

o and submitted to peer review journal.

It will be disseminated locally via the trust operational update and events, as well as more widely through national and international conference presentations. A summary of the work will be submitted for peer-reviewed publications. Participants wil be able to request a copy of the final study report.

### 9.2 Authorship Eligibility Guidelines

Authorship will take into account all persons involved in the study design, analysis and write up. This will be accurately reflected when any papers are submitted for peer-reviewed publication.

## Supporting information

Interview schedule

## Data Availability

All data produced in the present study are available upon reasonable request to the authors

## 11 Appendicies

### 11.1 Appendix 1-Required documentation

The CVs of members of the research team have been sent to the Research and Innovation Department.

