## Supplementary material for "DEFINE(prehospital): A protocol for a qualitative interview study to determine the pathway and response to the inclusion of frailty in pre-hospital trauma triage tools": Interview schedule

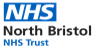

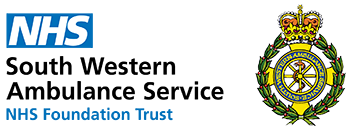

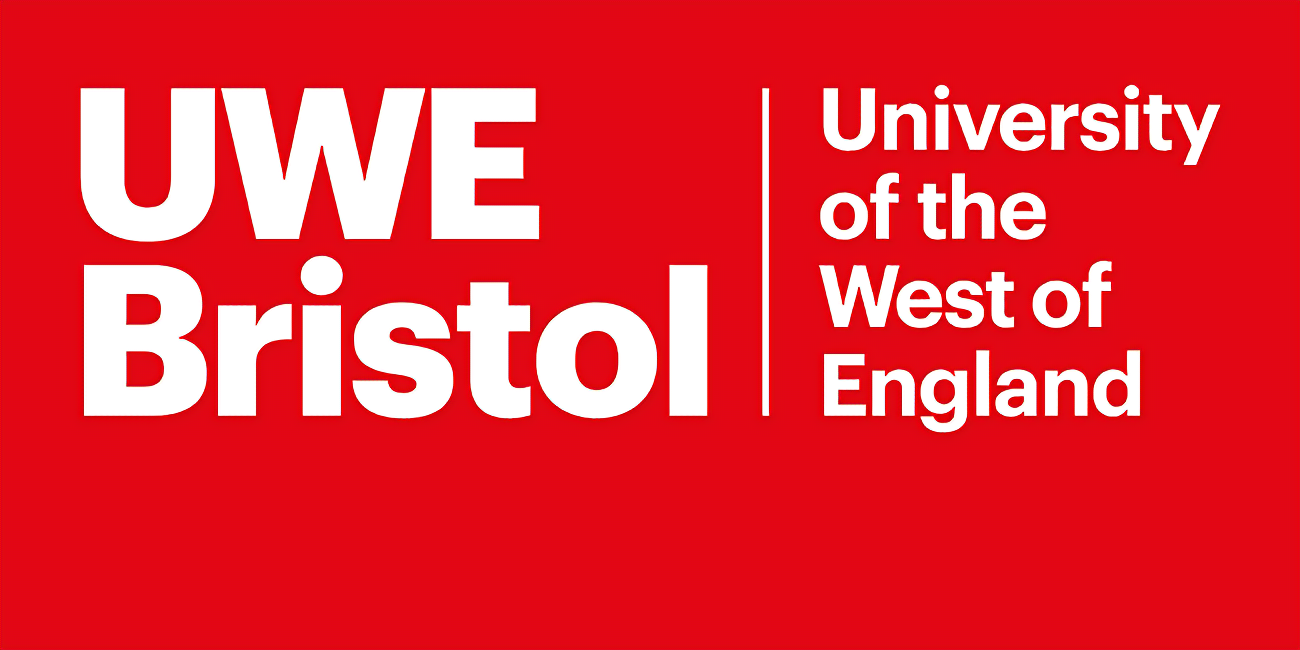

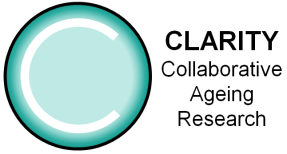

**DEFINE(prehospital): A qualitative interview study to determine the pathway and response to the inclusion of frailty in pre-hospital trauma triage tools**

*The pre-hospital frailty triage tool for SWASFT will be provided prior to interview with the below questions to consider. (SWASFT major trauma triage tool.pdf)*

Before our meeting, please have a look at the SWASFT major trauma triage tool, and in particular the central green box stating *Clinical Frailty (Rockwood) score of 5 or over*, and consider the following three questions:

1) Were you aware of that frailty is included in triage tool?
2) Do you know how frailty came to be included?
3) In your experience how has frailty been used in the triage tool?

**Interview Questions**

1) Background information

- Tell me your job title
- Where are you working most of the time?
- How long have been working in this role?

2) Pathways associated with the inclusion of a frailty assessment

- Did you have a chance to look at the SWASFT trauma triage tool I sent beforehand?
- Do you ever use the trauma triage tool in your job?
- Did you know that it included frailty?
- Do you know how frailty came to be included in the triage tool?
- Could you describe to me the pre-hospital trauma triage pathway and how frailty fits in.

3) Experiences of using frailty assessment in the pre-hospital setting

- In your experience is this what happens with frailty in pre-hospital environment?
- Have you ever discussed frailty when deciding where to take an injured person?
- What was the response of the person you spoke to about frailty?
- Do you think there is a difference between trauma units and MTCs in their response to frailty?

4) Views on the benefit and risks associated with frailty assessment

- Do you think frailty should be included in pre-hospital trauma triage?
- Have you experienced any benefits in assessing frailty pre-hospital?
- Have you experience any negatives in assessing frailty pre-hospital?
- In your opinion, is there anything that could make it easier to use frailty pre-hospital?
